# Comparison of CT-Derived Plaque Characteristic Index with CMR Perfusion for Ischemia Diagnosis in Stable CAD

**DOI:** 10.1101/2023.06.15.23291363

**Authors:** Wei-feng Guo, Hai-jia Xu, Yi-ge Lu, Guan-yu Qiao, Shan Yang, Shi-hai Zhao, Hang Jin, Neng Dai, Zhi-feng Yao, Jia-sheng Yin, Chen-guang Li, Wei He, Mengsu Zeng

## Abstract

**Background:** Coronary CT angiography (CCTA) and cardiac magnetic resonance (CMR) have been used to diagnose lesion-specific ischemia in patients with coronary artery disease (CAD).

**Objective:** The aim of this study was to investigate the diagnostic performance of CCTA-derived plaque characteristic index compared with myocardial blood flow (MBF) and myocardial perfusion reserve (MPR) derived from CMR perfusion in the assessment of lesion-specific ischemia.

**Methods:** Between October 2020 and March 2022, consecutive patients with suspected or known CAD, who were clinically referred for ICA were prospectively enrolled. All participants sequentially underwent CCTA and CMR and invasive fractional flow reserve (FFR) within 2 weeks. The diagnostic performance of CCTA-derived plaque characteristics, CMR perfusion-derived stress MBF, and MPR were compared. Lesions with FFR ≤ 0.80 were considered to be hemodynamically significant stenosis.

**Results:** Nighty-two patients with 141 vessels were included in this study. Plaque length, minimum luminal area, plaque area, percent area stenosis, total atheroma volume, vessel volume, lipid rich volume, spotty calcium, napkin-ring signs, stress MBF and MPR in flow-limiting stenosis group were significantly different from non-flow limiting group. The overall accuracy, sensitivity, specificity, PPV, and NPV of lesion-specific ischemia diagnosis were 61.0%, 55.3%, 63.1%, 35.6%, 79.3% for stress MBF, and 89.4%, 89.5%, 89.3%, 75.6%, 95.8% for MPR, meanwhile 82.3%, 79.0%, 84.5%, 65.2%, 91.6% for CCTA-Derived plaque characteristic index.

**Conclusion:** In our prospective study, CCTA-derived plaque characteristics and MPR derived from CMR performed well in diagnosing lesion-specific myocardial ischemia, and were significantly better than stress MBF in stable coronary artery disease.

## Introduction

Coronary artery disease (CAD) is a global problem and a leading cause of death(1). Myocardial ischemia causes permanent regional wall motion abnormalities, impairs cardiac function and negatively impacts the prognosis of patients with CAD(2). Therefore, the assessment of ischemia is a vital part of diagnosing stable CAD and guiding coronary revascularization(3).

In patients with stable CAD, invasive coronary angiography (ICA) with fractional flow reserve (FFR) measurement is considered to be the reference standard in the assessment of hemodynamically significant coronary stenosis(4), but clinically limited since it is invasive and expensive. Non-invasive modalities such as coronary CT angiography (CCTA) and cardiac magnetic resonance (CMR) imaging have gradually been recommended for the detection of clinically significant CAD(5). CCTA–derived(6) quantitative plaque characteristics provide valuable anatomic information with high sensitivity and negative predictive value for diagnosing obstructive CAD (7–10). In addition, quantitative CMR perfusion was able to rapidly quantify myocardial blood flow (MBF), validated by positron emission computed tomography(11), performed better than visual analysis. Myocardial perfusion reserve (MPR) derived from MBF has great potential for detecting myocardial ischemia(11–13).

Yet, to our knowledge, the direct comparison of diagnostic performance between CCTA-derived plaque characteristics and quantitative perfusion high-resolution CMR in identifying lesion-specific ischemia of stable CAD has not been investigated. Therefore, our study aims to investigate the diagnostic performance of plaque characteristics on CCTA compared with MBF and MPR on CMR in patients with stable CAD.

## Methods

### Patient Population and Study Design

From October 2020 to March 2022, a total of 110 patients with suspected or known CAD who were referred for clinically indicated ICA were prospectively recruited. Exclusion criteria included (1) Clinical unstable patients, (2) allergy to iodinated contrast material, (3) inability to undergo adenosine testing or CMR, (4) a glomerular filtration rate ≤45 ml/min, (5) a body mass index ≥40 kg/m2, (6) prior revascularization, (7) persistent atrial fibrillation, (8) pregnancy or (9) life expectancy < 2 years caused by noncardiac disease, (9) not eligible for FFR measurement as extremely tortuous, calcified coronary vessels or chronic total occlusion. Consequently, the study population included subjects who had between 30% and 90% stenosis of ≥1 major epicardial vessels by ICA, and whose invasive FFR measured in that vessel within 2 weeks of coronary CTA and quantitative stress perfusion CMR. The decision on revascularization was guided by invasive FFR in stenosis ranging from 30% to 90% and was made at the discretion of the operator or the Heart Team. The study flow chart is shown in Figure 1. The study was approved by the hospital institutional review board. All participating patients gave their informed consent for the study.

**Figure 1.**
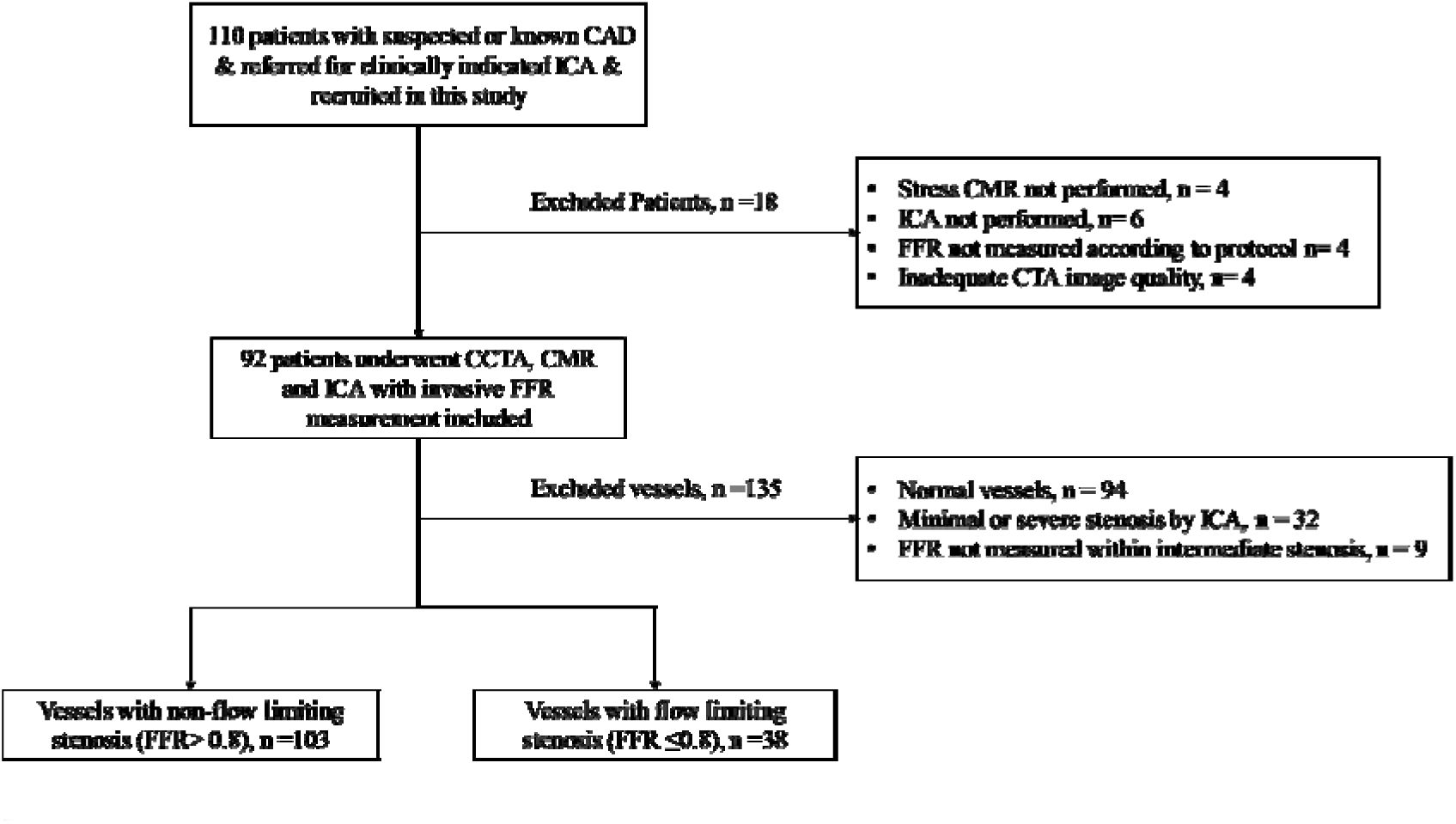
Study flow chart.

### Coronary Computed Tomography Angiography

Third-generation dual-source CT (DSCT) systems ((SOMATOM Force, Siemens, Forchheim, Germany)) were used for image acquisition. Sublingual nitroglycerine was administered to all patients and metoprolol only if necessary to lower heart rate to 60 bpm or below before cardiac CT scanning. The prospectively ECG-triggered high-pitch spiral scan with a tube voltage of 120 kV was used for an initial non-contrast-enhanced calcium scoring scan. CCTA acquisitions were performed prospectively using the third-generation DSCT with the following parameters: tube voltage automatically selected using an automated tube-voltage selection algorithm (CARE kV, Siemens Healthineers, Germany), 192×0.6 mm collimation, 0.25 s rotation time, tube current determined using an automatic exposure control system. The reference quality is 300 or 350 mAs/rotation with 120kV. CCTA axial image reconstruction was using the smooth kernel (Bv40) and iterative reconstruction technique (strength 3, ADMIRE, Siemens) with a thickness and increments of 0.6 mm and 0.3 mm.

### Analysis of CCTA data and plaque quantification

CCTA assessments were performed on the color-coded semi-automatic plaque analysis software (Coronary Plaque Analysis, version 2.0, Siemens Healthineers) by skillful CT cardiologists, who had ≥10 years of experience in CCTA interpretation and were blinded to other analyses. We analyzed one target lesion per vessel with a 30% to 90% stenosis where invasive FFR was performed. When a vessel presented with more than one separate lesions, the severest lesion was included into the study. As for diffuse lesions, we analyzed the segment across the lesion, from the proximal normal segment to distal normal segment(14). At the point free of atherosclerotic plaque, the average diameter and area of the regions proximal and distal to the lesion of interest were measured as a reference. The quantitative parameters of plaque analysis were measured automatically by the software (Figure 2) and all the parameters used in our study were defined as follows: 1) plaque length or area refers to the length or area per lesion, 2) MLA was defined as the minimum luminal area per vessel(15), 3) Percent area stenosis (PAS in %) was defined as the ratio of the plaque area over the reference area of the vessel (16), 4)The contours of the lumen and external elastic membrane were manually traced at every 0.5-mm cross-sectional frame of the vessel using the software, generating volumetric data to describe lumen volume and vessel volume. Total atheroma volume (TAV) was calculated as vessel volume minus lumen volume(17). 5) The lipid plaque ratio was defined as the ratio of the lipid plaque volume to the total atheroma volume. 6) Spotty calcium was assessed visually as calcification with a length above 3mm as well as covering <90° of the circumference of the blood vessel(7). 7) Remodeling index was measured as the ratio of the vessel area of the lesion over the proximal luminal area of the reference area(18), and positive remodeling was defined as remodeling index ≥1.1(19), 8) The presence of the napkin-ring sign was assessed as a low attenuation necrotic core circumscribed by an area of higher attenuation(7), 9) low attenuation plaque was defined as the presence of plaque with attenuated CT value.

**Figure 2.**
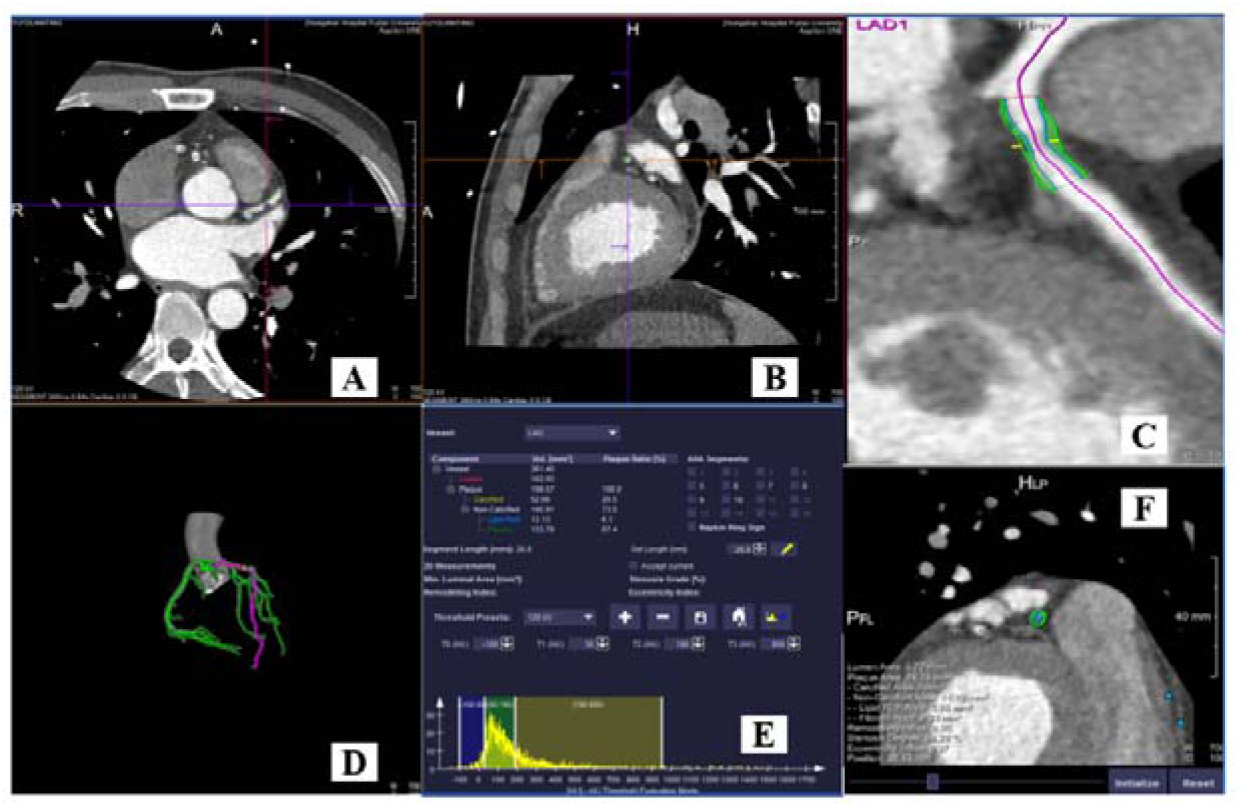
Case example of CCTA-derived plaque characteristic index analysis. Representative patient who had underwent CCTA for suspected CAD. (A, B) The original image data of the patient. (C, D) Reconstruction image and model. (E) Color-coded semiautomatic plaque quantification of the lesion shows plaque length (18.9mm), plaque area (10.47), MLA (2.9) and etc. (F) 2D plaque characteristics analysis shows the lesion without any spotty calcium, positive remodeling, napkin-ring signs and low attenuation plaque.

### Stress Cardiac Magnetic Resonance Perfusion Protocol

All patients were asked to refrain from caffeine-containing foods and beverages for 24h before the scan. All scans were performed on a 3-T scanner (MAGNETOM Prisma; Siemens Healthcare, Erlangen, Germany). 2-, 3-, and 4-chamber long-axis cine images were acquired as well as contiguous short-axis slices covering the left ventricle. To induce the maximum hyperemia, intravenous adenosine triphosphate (ATP) of 140 ug/kg/min was infused, but if there weren’t any symptoms after ATP infusion including no ≥10 beats per minute heart rate increase and no ≥10 mmHg blood pressure drop(20), then increased to 210 ug/kg/min. After 4 min of ATP infusion, stress perfusion data were acquired with the following parameters: repetition time (TR) of 2.9ms, echo time (TE) of 1.0ms, spatial resolution of 1.9×2.5×8.0 mm^3^, and temporal resolution of 110ms. The arterial input function (AIF) was corrected using a dual-bolus gadobutrol (Gadovist; Bayer, Berlin, Germany) contrast agent administration. 5 baseline frames were acquired before the injection of AIF bolus of gadobutrol (small dose, 0.0075mmol/kg), then injection of subsequent 25ml saline at 2ml/s was started, followed by the acquisition of consistent stress AIF of 60 frames without breath holding. Stress myocardial perfusion images were acquired subsequently with the injection of the main bolus of gadobutrol (big dose, 0.075mmol/kg), and other operations were the same as AIF acquisition. Resting perfusion imaging was performed 15 min after the stress, using the same protocol. To correct the intensity inhomogeneities of surface coils, the weighted proton density acquisition was performed before stress and rest acquisitions.

### Analysis of Cardiac Magnetic Resonance

CMR was analyzed offline using commercially available software (CVI42, v.5.13, Circle Cardiovascular Imaging, Calgary, Alberta, Canada). All CMR images analysis were conducted by skillful radiologists with >10 years of expertise in CMR perfusion imaging and quantitative analysis was carried out by one of the experts in a blinded fashion. Before the quantitative analysis, the perfusion images were motion corrected according to published methods(21), and quantitative perfusion analysis was performed by Fermi-constrained deconvolution as the previously described methods(22). The division of the myocardial border was automatedly and manually corrected when required, to exclude coronary arteries and obvious image artifacts. Average MBF was quantified in ml/min/g and was assessed using the American Heart Association (AHA) modified 17-segment model (the apical segment was excluded)(20). Myocardial perfusion reserve (MPR) was defined as the ratio of stress MBF to rest MBF. Stress MBF and MPR were measured and analyzed subsequently (Figure 3). Observers would review the images together to reach a consensus in case of disagreement.

**Figure 3.**
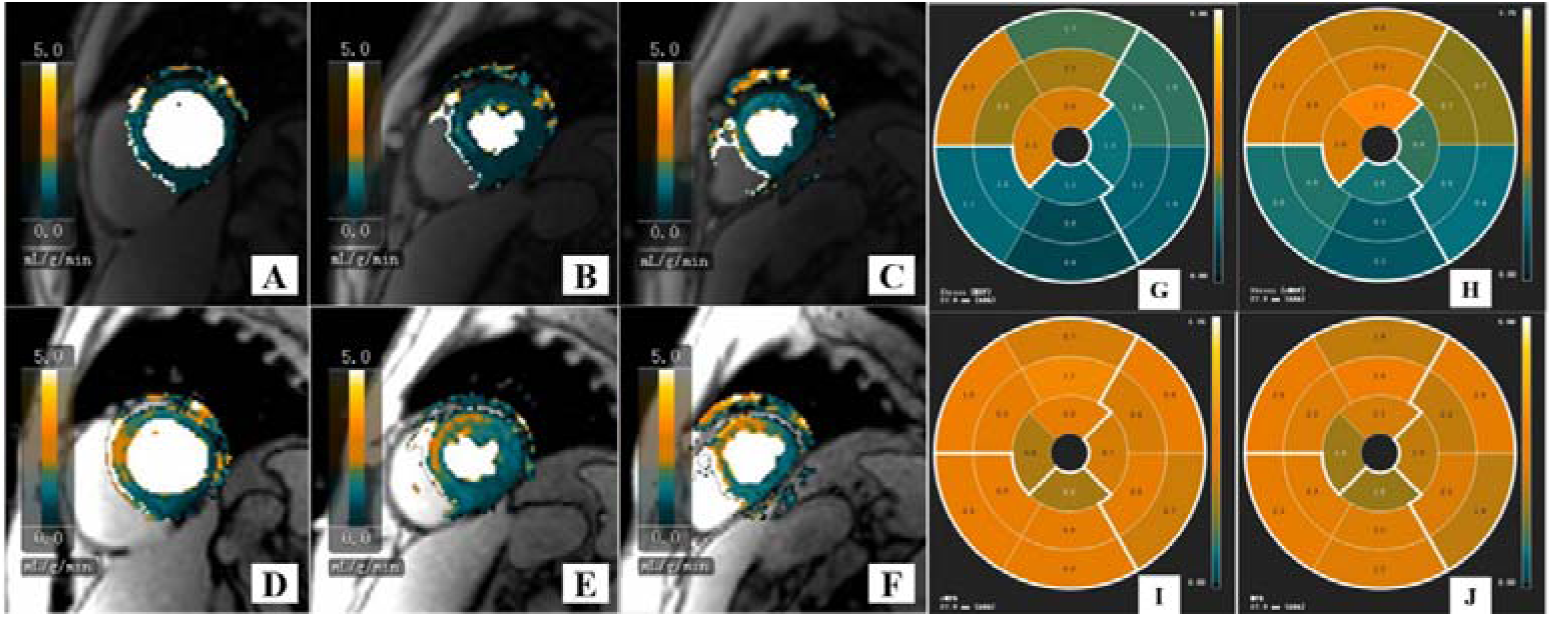
Case example of quantitative CMR perfusion analysis. Representative patients with chest pain suspected of CAD. CCTA showed moderate stenosis of LAD. (A-E) Cardiac magnetic resonance (CMR) imaging of the basal left ventricular (A, D), midventricular (B, E), and apical (C, F) slices are shown during hyperemic (D-F) and resting (A-C) conditions. (G-J) The 17-segment Bulls-eye diagrams showing the distribution of CMR perfusion of stress MBF (G, H) and MPR (I, J) according to LAD.

### Invasive Coronary Angiography and Fractional Flow Reserve Measurement

ICA was performed according to current guidelines(23). FFR measurements were conducted on each coronary segment by experienced interventional cardiologists in a blinded fashion. A pressure wire (Aeris; St. Jude Medical, St. Paul, Minn) was used to obtain FFR during continuous infusion of intravenous ATP (140ug/kg/min), while the pressure sensor was positioned distal to the lesion where ICA determined 30% to 90% coronary stenosis in the main branch coronary arteries (greater or equal to 2 mm). Hyperemic proximal aortic pressure (Pa) and distal arterial (Pd) pressure were measured during induced hyperemia, and FFR was calculated automatically as the ratio of mean Pd to mean Pa. Vessel would be diagnosed as flow-limiting significant if FFR value ≤0.80.

### Statistical Analysis

Statistical analysis was carried out by SPSS 20.0 and MedCalc software. Normal distribution was assessed by use of the Kolmogorov-Smirnov (K-S) test. Continuous variables were displayed as mean ± standard deviation (SD) and compared by Student *t* test or Student *t’* test as appropriate, or expressed as median with an interquartile range (IQR) and compared by Mann-Whitney *U* test when not normally distributed. Categorical variables were shown as numbers and compared using Pearson chi-squared test. Parameters of CCTA-derived plaque characteristics that significantly differed between the flow-limiting group and non-flow-limiting group were then included into the univariate and subsequent forward stepwise multivariate logistic regression analysis. CCTA-derived plaque characteristics logistic regression model was created based on the independent predictors of lesion-specific ischemia. Receiver operating characteristic (ROC) analysis was used to analyze the diagnostic performance of CCTA-derived quantitative plaque characteristics, stress-MBF, and MPR. The Youden index derived from the ROC curve analysis was used to determine the optimal cutoff. The area under the ROC curve (AUC) was measured and compared using Delong’s test to evaluate the discriminatory power as well as the sensitivity, specificity, positive predictive value (PPV), and negative predictive value (NPV) with a 95% confidence interval (CI). Two-tailed p values <□0.05 were considered significant.

## Results

### Patient characteristics

Of 110 patients enrolled, 106 patients underwent all noninvasive imaging within 2 weeks before ICA, and 14 patients were excluded due to not undergo CMR (n=4) or ICA (n=6) or invasive FFR (n=4). 4 patients were excluded for inadequate CCTA image quality. Therefore, 92 patients comprised the final study population, of whom 141 vessels with stenosis ranged between 30% and 90% underwent all tests (Figure 1). Detailed patient baseline characteristics were presented in Table 1. 38 lesions (27.0%) demonstrated lesion-specific ischemia (FFR ≤ 0.80) were divided into the flow-limiting group. The remaining 103 lesions (73.0%) were not hemodynamically significant stenosis (FFR > 0.80) and were divided into the non-flow-limiting group (Table 1).

**Table 1.**
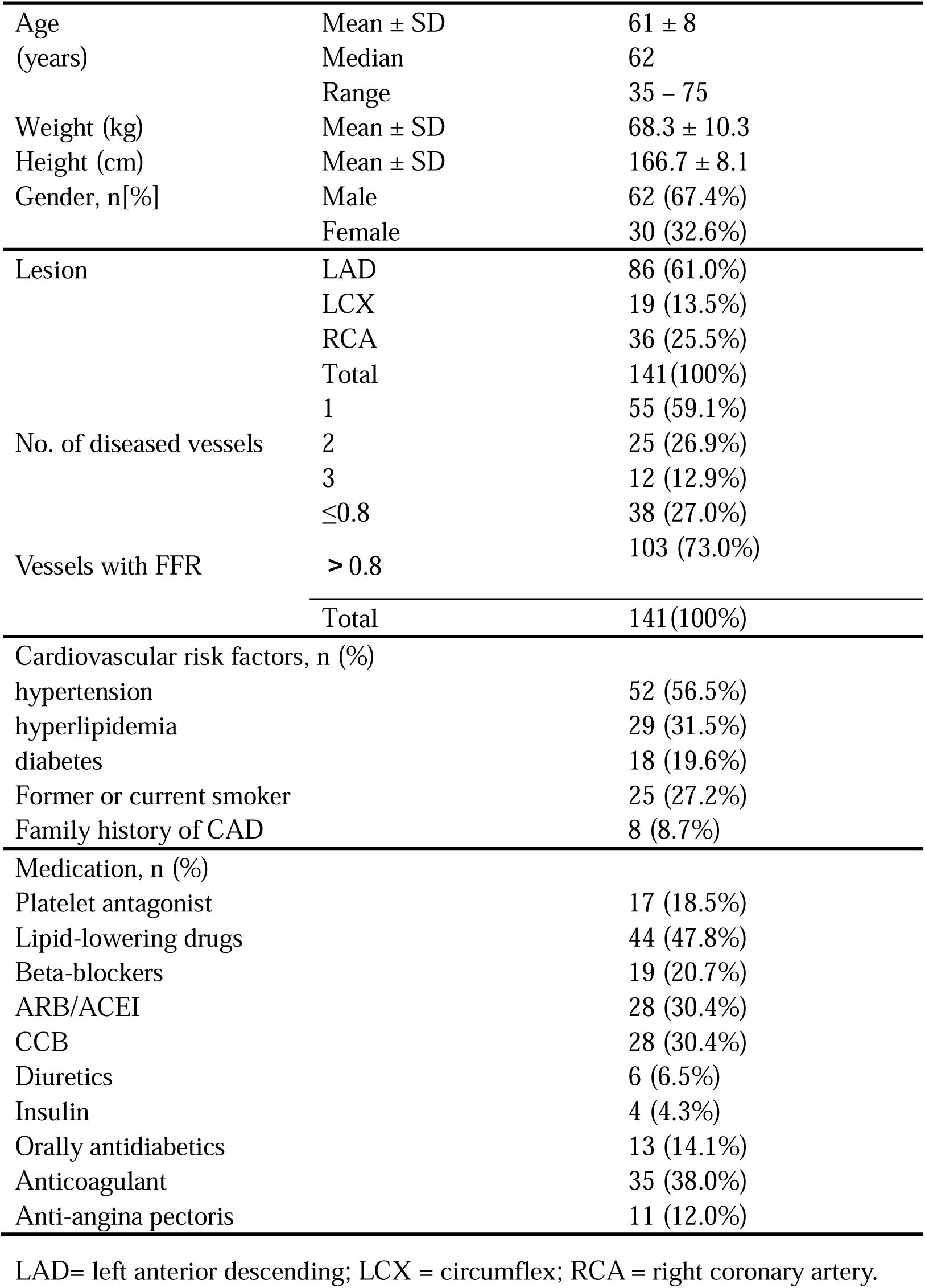
Patient demographics. Total patient cohort (n =92)

### CCTA-derived plaque characteristics, MBF and MPR derived from CMR perfusion

All CCTA-derived plaque characteristics and myocardial perfusion data derived from CMR were summarized in Table 2. Plaque length (27.2 ± 11.6 vs 37.1 ± 15.3mm, p<0.001), plaque area (8.1 ± 3.3 vs 10.1 ±4.3*mm*^2^, p=0.007), percent area stenosis (PAS, 53.4 ± 16.3 vs 74.4 ± 15.3, p<0.001), total atheroma volume (TAV, 171.6 ± 110.4 vs 270.1 ± 164.2*mm*^2^, p<0.001), vessel volume (360.5 ± 202.4 vs 441.9 ± 235.7*mm*^3^, p=0.045), lipid rich volume (median 10.3 vs 21.7*mm*^3^, p=0.01), spotty calcium (p<0.001), and napkin-ring signs (p<0.001) in flow-limiting stenosis group were significantly higher than non-flow-limiting stenosis group, while minimum luminal area (MLA, median 3.4 vs 1.5*mm*^2^, p<0.001), stress MBF(2.8 ± 0.7 vs 2.4 ± 0.7mL/(g·min), p=0.007) and MPR (myocardial perfusion ratio, 2.6 ± 0.9 vs 1.6 ± 0.4, p<0.001) in flow-limiting stenosis group were significantly lower than non-flow-limiting stenosis group. Lipid plaque ratio, positive remodeling, and low attenuation plaque showed no significant differences between flow-limiting group and non-flow-limiting stenosis group (Table 2).

**Table 2.**
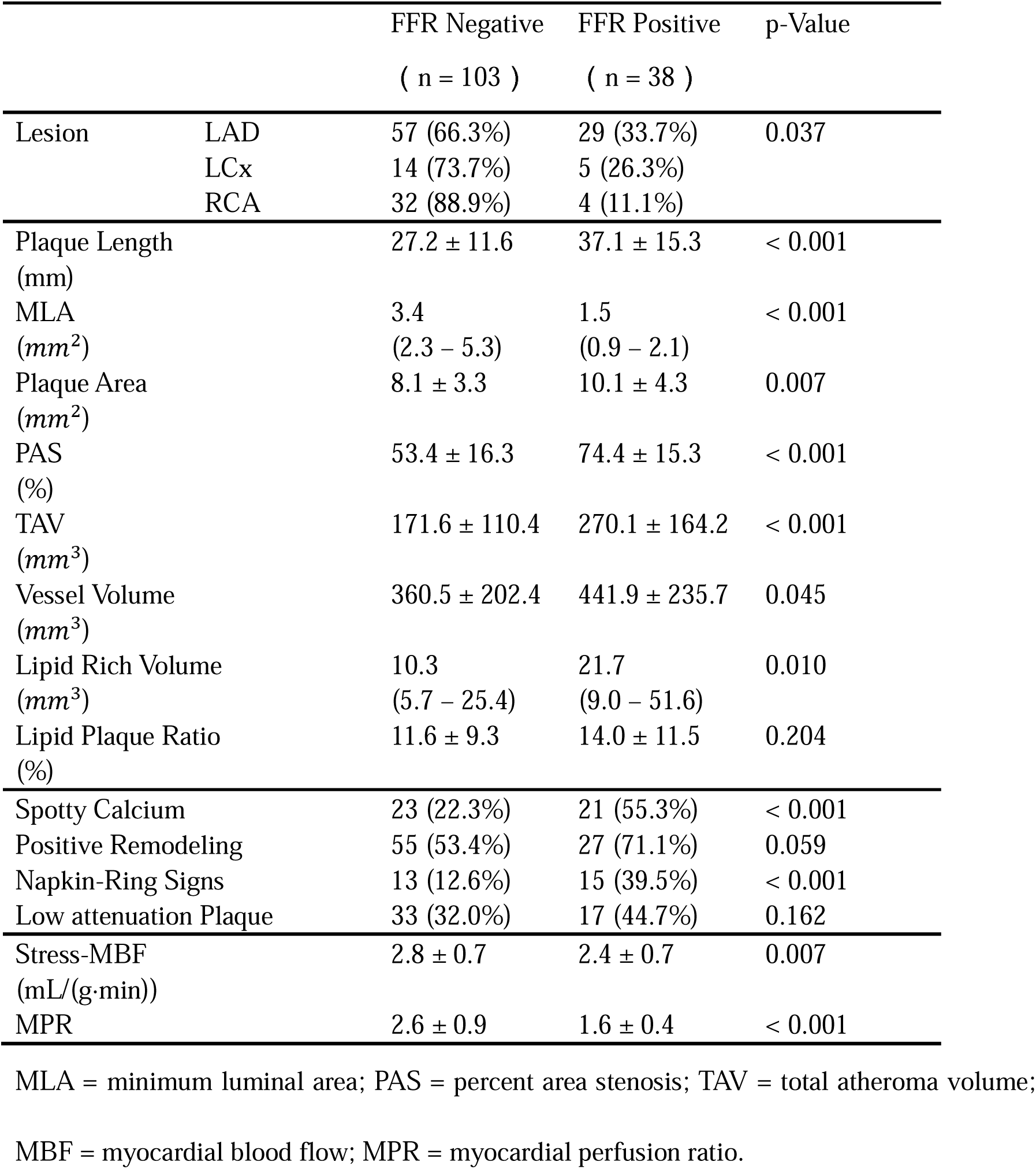
Plaque characteristics and CMR data.

### The univariate and multivariate logistic regression analyses of plaque characteristics for lesion-specific ischemia

Variables that significantly different between the flow-limiting and non-flow-limiting stenosis groups were included in the univariate logistic regression analyses, and independent association were found for plaque length (OR, 1.059, p<0.001), MLA (OR, 0.305, p<0.001), plaque area (OR, 1.142, p=0.009), PAS (OR, 1.095, p<0.001), TAV (OR, 1.006, p<0.001), vessel volume (OR, 1.002, p=0.049), lipid rich volume (OR, 1.016, p=0.005), spotty calcium (OR, 0.233, p<0.001) and napkin-ring signs (OR, 0.221, p=0.001). Variables that significantly differed in the univariate logistic regression analyses were then included in the multivariate analyses, and MLA (OR, 0.284, p<0.001) and TAV (OR, 1.006, p=0.003) were found to be the significant independent predictors of lesion-specific ischemia (Table 3). Multivariate logistic regression analysis showed that the correlation coefficients of lesion-specific ischemia with plaque characteristics with MLA and TAV were -1.25891 and 0.005841, respectively (P<0.001, P=0.003, respectively), and the regression equation of CCTA-derived plaque characteristic index for predicting probability P was *P* = 1/[1 + *e*^-(-1.258591xMLA+0.005841xTAV+0.803637)^].

**Table 3.**
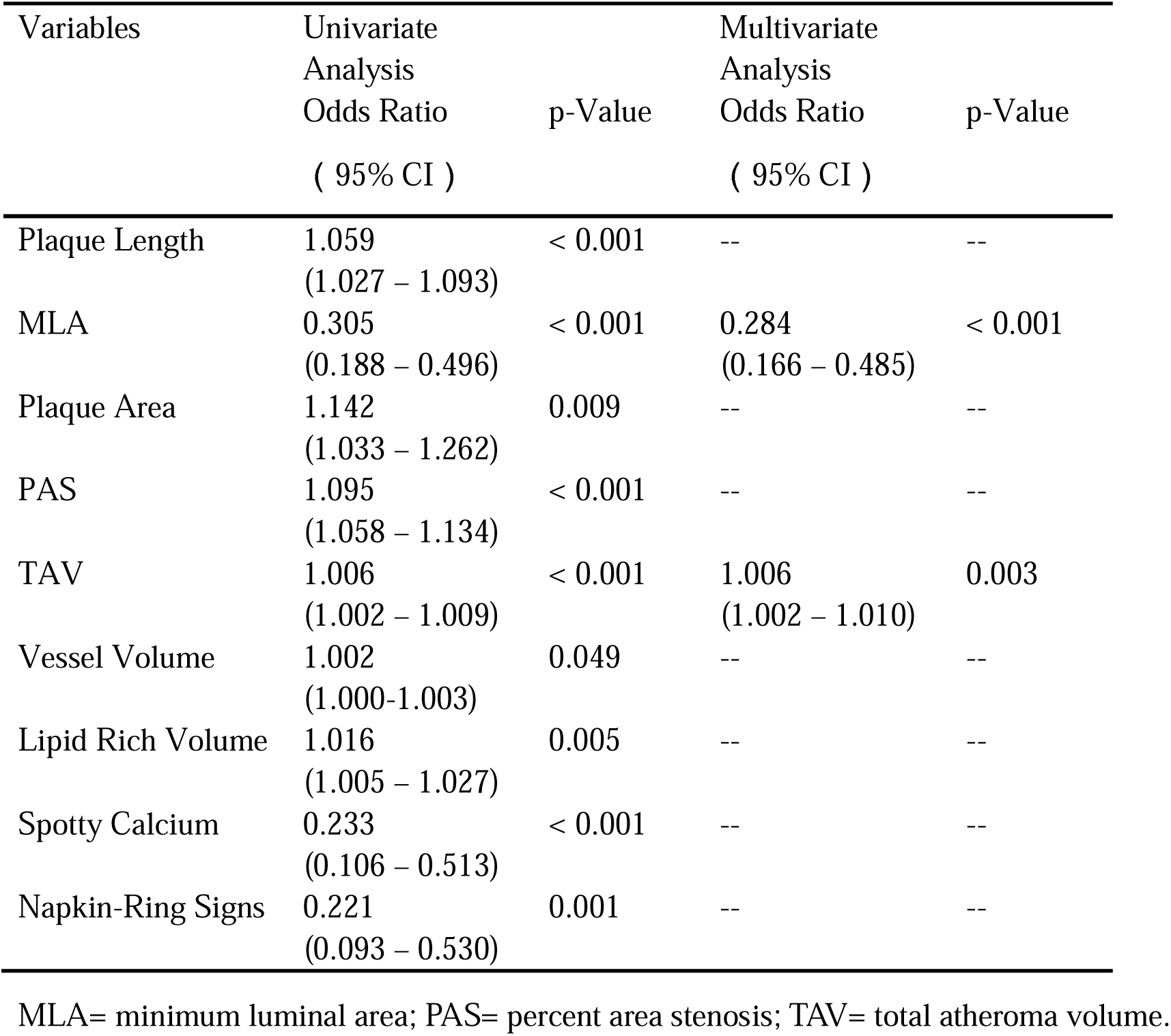
Univariate and multivariate logistic regression analysis of plaque characteristics.

### The diagnostic performance of CCTA-derived plaque characteristic index, stress-MBF, and MPR for myocardial ischemia

The optima cut-off value based on Youden’s index for MLA and TAV were 2.25 and 161.72. And the diagnostic performance for MLA and TAV in detecting lesion-specific ischemia were shown in the Table S1. The optimal cut-off value based on Youden’s index for stress-MBF, MPR and plaque characteristic index were 2.61, 1.82, and 0.368. The area under the ROC curve (AUC) of MPR (0.921) showed no statistical discrepancies with plaque characteristic index (0.885, p=0.38), while both were significantly higher than stress MBF (0.638, p<0.001, figure 4). The overall diagnostic accuracy, sensitivity, specificity, PPV, and NPV of stress MBF for detecting lesion-specific ischemia were 61.0%, 55.3%, 63.1%, 35.6%, 79.3%, and MPR was 89.4%, 89.5%, 89.3%, 75.6%, and 95.8%, meanwhile plaque characteristic index was 82.3%, 79.0%, 84.5%, 65.2%, and 91.6% (Table 4).

**Figure 4.**
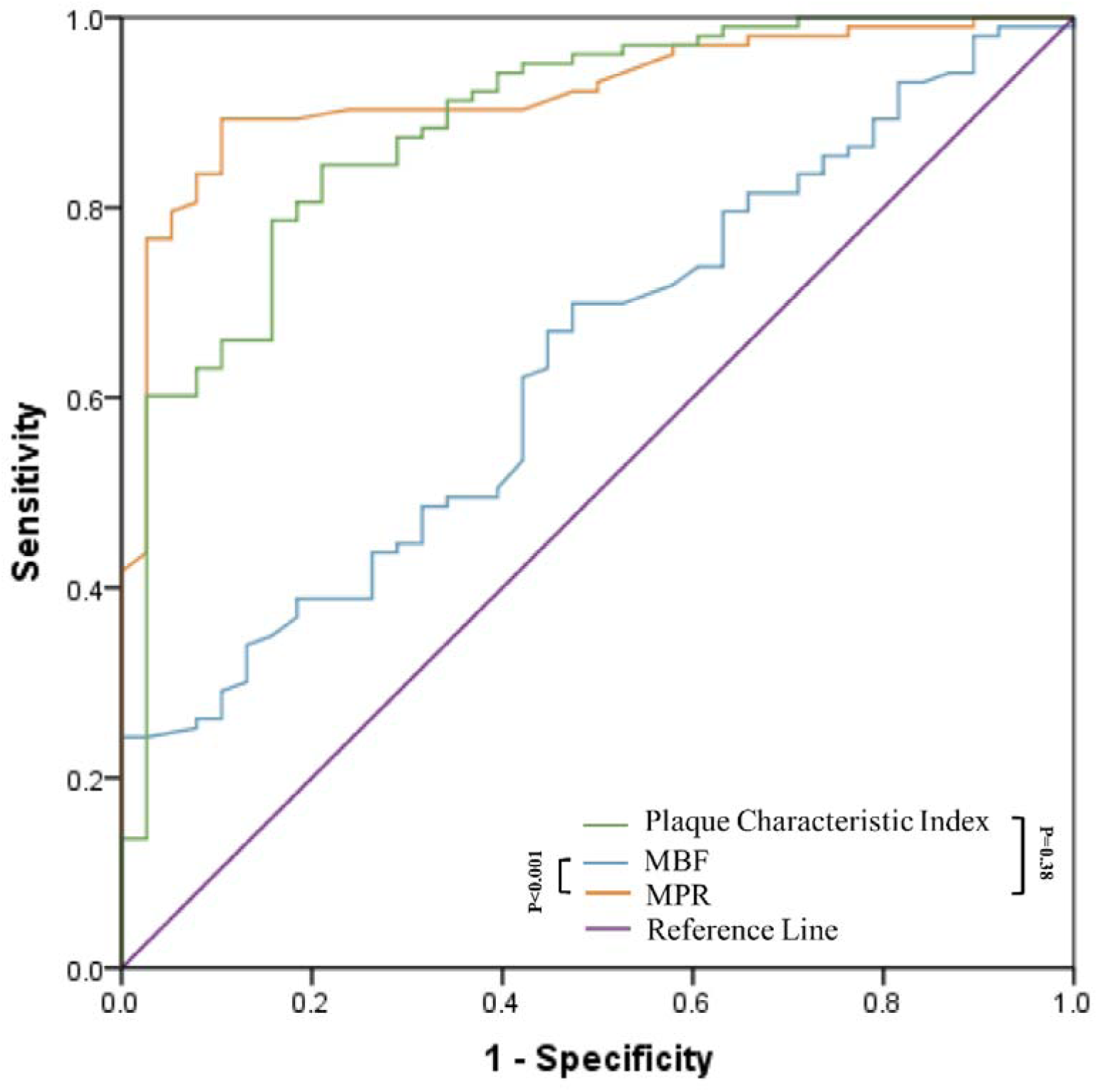
Diagnostic performance of stress MBF, MPR, and Plaque Characteristic Index for the prediction of lesion-specific ischemia. The ROC analysis demonstrated that AUC of MPR (0.921, 95% CI: 0.863–0.959) and plaque characteristic index (0.885, 95% CI: 0.821–0.933, p=0.38) were higher than stress MBF (0.638, 95% CI: 0.553–0.718). MBF= myocardial blood flow; MPR= myocardial perfusion reserve.

**Table 4.**
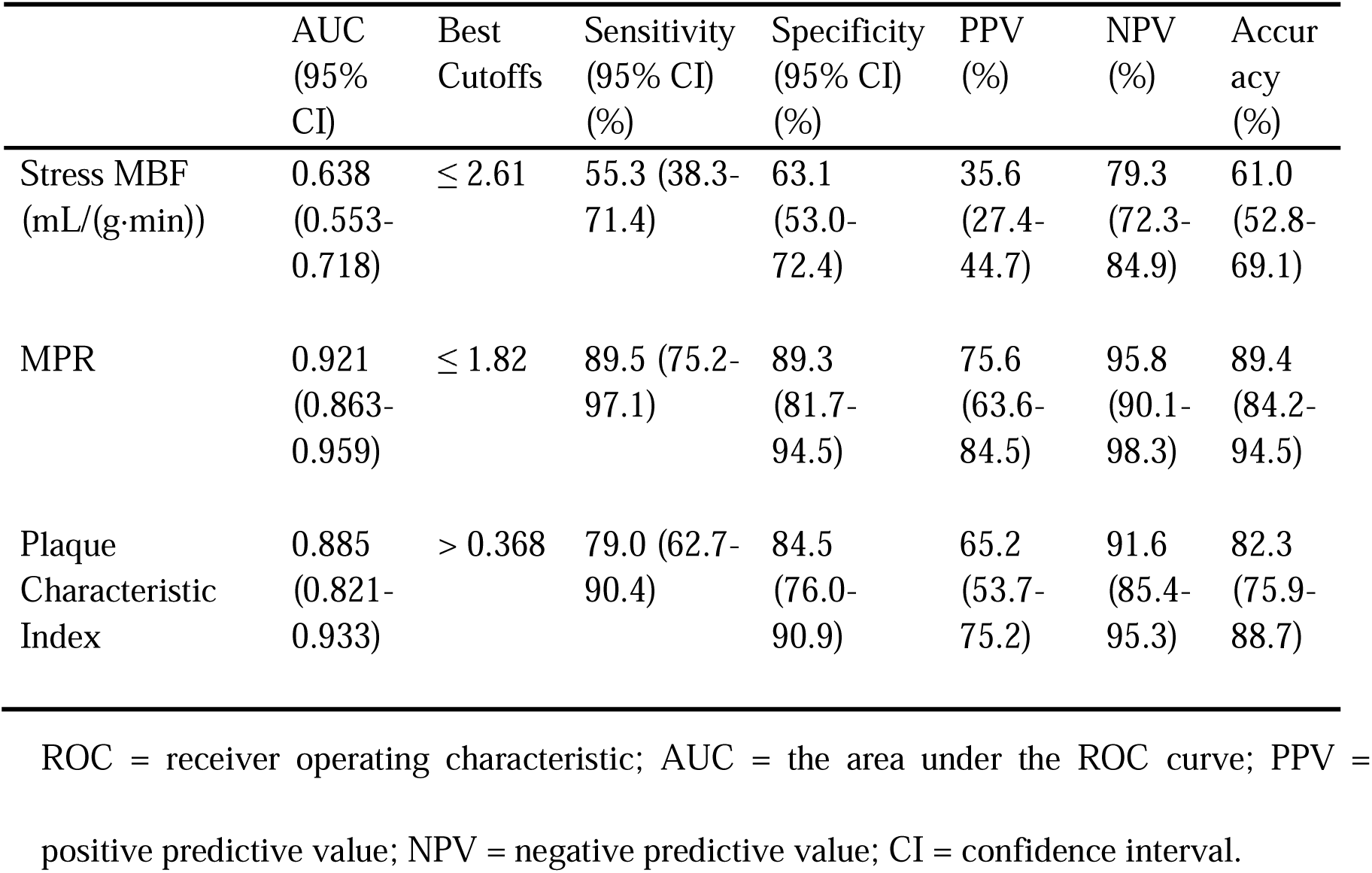
Diagnostic performance of stress MBF, MPR, and Plaque Characteristic Index for detecting lesion-specific ischemia.

## Discussion

We prospectively compared the performance of plaque characteristic index, CMR-derived stress MBF and MPR in diagnosing lesion-specific ischemia, and it is the first time, to our knowledge (Central illustration). Our results demonstrated that some CCTA-derived characteristics (plaque length, MLA, plaque area, PAS, TAV, etc.) and quantitative CMR perfusion were statistically different between patients with hemodynamically significant and non-hemodynamically significant lesion. And the diagnostic efficiency of plaque characteristics and MPR were both well and superior to stress MBF. MLA and TAV were independent predictors for lesion-specific ischemia, which showed great diagnostic performance, showing no statistical discrepancies with MPR derived from CMR. Both of them performed better than stress-MBF in the diagnosis of lesion-specific ischemia.

Our studies reported that CCTA-derived plaque characteristics including plaque length, MLA, plaque area, percent area stenosis, total atheroma volume, vessel volume, lipid rich volume, spotty calcium, and napkin-ring signs were significantly different in the flow-limiting group from the non-flow limiting group, indicating the connection between quantitative plaque characteristics and hemodynamically stenosis, which was consistent with previous studies(9,15). Besides, MLA and PAV (percent atheroma volume) were independent predictors of myocardial ischemia according to the multivariate logistic regression, and the report by Seokhun Yang et al. (16) provided evidence to support our results that MLA and PAV were selected as the top two relevant features for myocardial ischemia in their study, by use of the Buruta algorithm and hierarchical clustering.

In the report by Sadako Motoyama et al.(24), positive remodeling and low attenuation plaque were considered to be the characteristics of high-risk plaque and an independent predictor of ACS (acute coronary syndrome), however, our study demonstrated that they had no significant differences between flow-limiting and non-flow-limiting stenosis groups, and were less informative for detecting myocardial ischemia in stable CAD.

In previous studies(11,12), quantitative analysis of high-resolution CMR was also considered to be a reliable method of detecting lesion-specific ischemia, and MPR derived from CMR has already been used to diagnose hemodynamically significant stenosis. In the study by Timothy Lockie et al. (11), the threshold of MPR was determined as 1.58 (AUC 0.92), while it was 1.82 (AUC 0.921) in our study, probably because the reference standard was FFR<0.75 in their study, while FFR≤0.80 was used in ours, therefore, it is understandable that the optimal threshold of MPR in their study was lower than ours. Therefore, more clinical studies are needed to find out a reliable threshold of MPR. Additionally, we also found that there were no statistical discrepancies between MPR based on CMR (AUC 0.921) and CCTA-derived plaque characteristics (AUC 0.885). The diagnostic performance of MPR and plaque characteristic index were both significantly higher than stress-MBF (AUC 0.638, p<0.001) and perform excellently in diagnosing myocardial ischemia in stable CAD as noninvasive modalities. Even though the diagnostic accuracy, sensitivity, specificity, PPV, and NPV of MPR were slightly higher than quantitative plaque characteristics, both of which can help reduce unnecessary invasive coronary angiography and FFR measurement on the diagnosis of myocardial ischemia.

## Limitation

Firstly, it is a single-center study with a small sample of subjects and vessels. Larger multicenter, well powered prospective studies are required to further establish the diagnostic performance of CCTA-derived plaque characteristics, CMR perfusion-derived MBF and MPR. The vessels with diameter stenosis <30% or >90% were not assessed because performing physiological assessments in such lesions was unnecessary. A total of 18 patients were excluded, which may incur a selection bias. Secondly, we haven’t investigated the diagnostic efficiency of MBF at rest, however generally at peak hyperemia the variability of resting vascular tone and hemodynamics will be eliminated(25) and the maximum myocardial blood flow will be achieved. Additionally, in our study, we chose FFR as the reference standard, rather than revascularization, which refer to clinical information such as symptoms and angiography apart from FFR(3). Finally, the CCTA-Derived plaque characteristic index needs to validate on a new dataset, and a larger multi-center study is needed to determine the optimal cutoff of plaque characteristics and MPR for the subsequent clinical application.

## Conclusion

In our prospective study, CCTA-derived plaque characteristics and MPR based on CMR performed well in diagnosing lesion-specific myocardial ischemia, and were significantly better than stress MBF.

## Supporting information

Manuscript marked version

Responses to Reviewer 2

Reply to the editor and reviewers

Response to editor

Responses to Reviewer 1

## Data Availability

All data produced in the present study are available upon reasonable request to the authors

## Acknowledgments

None

## Ethical approval

Ethics committee of Zhongshan Hospital, Fudan University, Shanghai, China, gave ethical approval for this work.

## Non-standard Abbreviations and Acronyms

CCTA: coronary CT angiography
MBF: myocardial blood flow
CMR: cardiac magnetic resonance
FFR: fractional flow reserve
MLA: minimum luminal area
PAS: percent area stenosis
TAV: total atheroma volume
MPR: myocardial perfusion reserve
CAD: coronary artery disease
ICA: invasive coronary angiography
DSCT: third-generation dual-source CT
ATP: intravenous adenosine triphosphate
ROC: receiver operating characteristic
AUC: the area under the ROC curve
PPV: positive predictive value
NPV: negative predictive value
CI: confidence interval
LAD: left anterior descending
LCX: circumflex
RCA: right coronary artery

---

**Figure 1.**
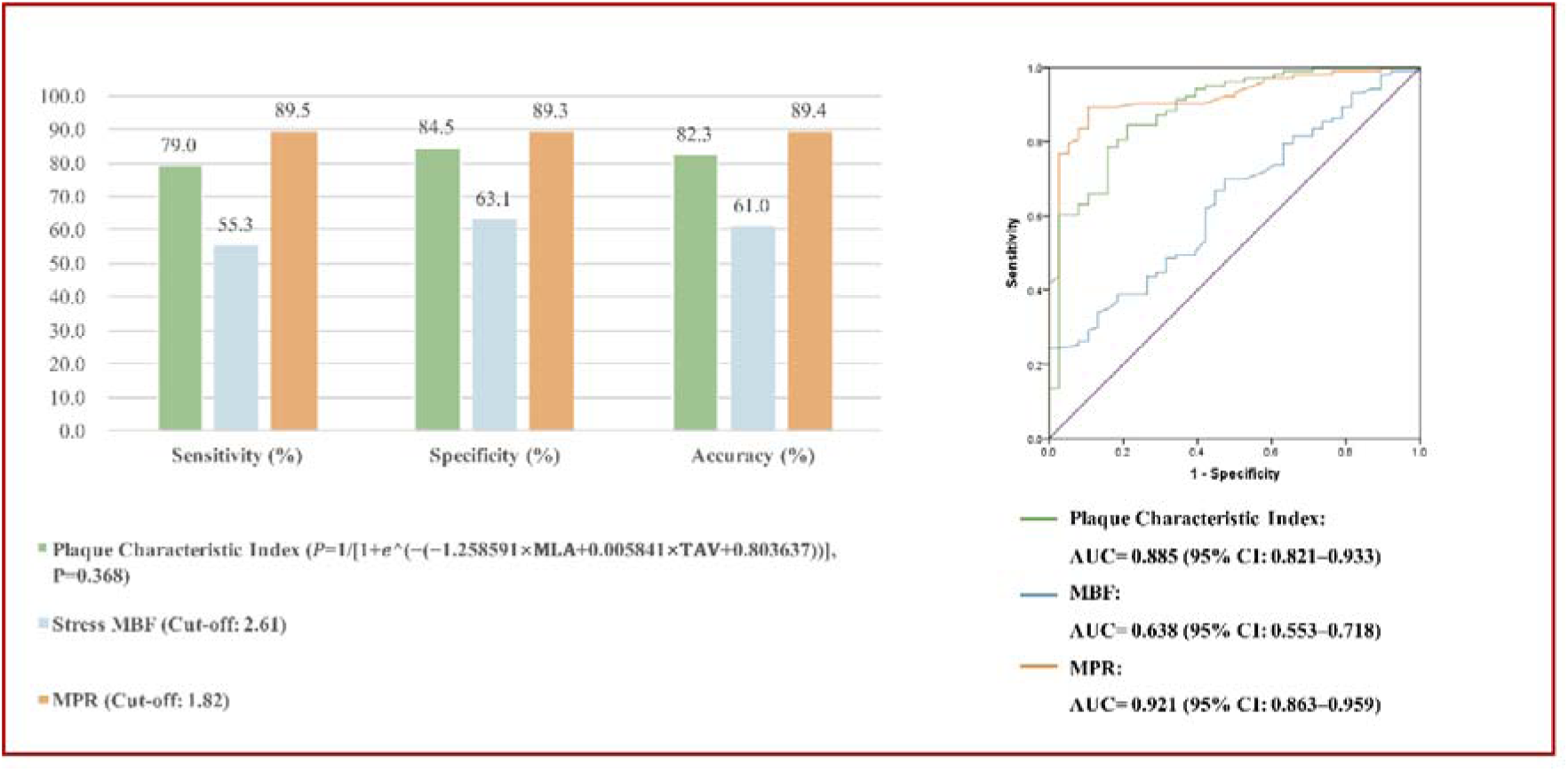
Central illustration. Diagnostic Performance of Plaque Characteristic Index derived from Coronary Computed Tomography Angiography, Stress Myocardial Blood Flow and Myocardial Perfusion Reserve to Identify Hemodynamically Significant Stenosis in Patients with Stable Coronary Artery Disease. The receiver-operating characteristic curve and corresponding area under the curve describing the diagnostic performance of plaque characteristic index derived from Coronary Computed Tomography Angiography (CCTA), stress myocardial blood flow (MBF) and myocardial perfusion reserve (MPR) to identify hemodynamically significant stenosis as defined by invasive coronary angiography (ICA) with fractional flow reserve (FFR) at the vessel level.

