## Supplementary material for "Comparison of CT-Derived Plaque Characteristic Index with CMR Perfusion for Ischemia Diagnosis in Stable CAD": Manuscript marked version

**Short title:** CCTA and MPR in myocardial ischemia

**Word count:** 4930

**Abstract**

**Background** Coronary CT angiography (CCTA) and cardiac magnetic resonance (CMR) have been used to diagnose lesion-specific ischemia in patients with coronary artery disease (CAD).

**Objective** The aim of this study was to investigate the diagnostic performance of CCTA-derived plaque characteristic index compared with myocardial blood flow (MBF) and myocardial perfusion reserve (MPR) derived from CMR perfusion in the assessment of lesion-specific ischemia.

**Methods** Between October 2020 and March 2022, consecutive patients with suspected or known CAD, who were clinically referred for ICA were prospectively enrolled. All participants sequentially underwent CCTA and CMR and invasive fractional flow reserve (FFR) within 2 weeks. The diagnostic performance of CCTA-derived plaque characteristics, CMR perfusion-derived stress MBF, and MPR were compared. Lesions with FFR ≤ 0.80 were considered to be hemodynamically significant stenosis.

**Results** Nighty-two patients with 141 vessels were included in this study. Plaque length, minimum luminal area, plaque area, percent area stenosis, total atheroma volume, vessel volume, lipid rich volume, spotty calcium, napkin-ring signs, stress MBF and MPR in flow-limiting stenosis group were significantly different from non-flow limiting group. The overall accuracy, sensitivity, specificity, PPV, and NPV of lesion-specific ischemia diagnosis were 61.0%, 55.3%, 63.1%, 35.6%, 79.3% for stress MBF, and 89.4%, 89.5%, 89.3%, 75.6%, 95.8% for MPR, meanwhile 82.3%, 79.0%, 84.5%, 65.2%, 91.6% for CCTA-Derived plaque characteristic index.

**Conclusion** In our prospective study, CCTA-derived plaque characteristics and MPR derived from CMR performed well in diagnosing lesion-specific myocardial ischemia, and were significantly better than stress MBF in stable coronary artery disease.

**Keywords coronary** CTA, CMR, CAD, Diagnostic Performance

**Non-standard Abbreviations and Acronyms**

**CCTA** = coronary CT angiography

**MBF** = myocardial blood flow

**CMR** = cardiac magnetic resonance

**FFR** = fractional flow reserve

**Acknowledgments**

None

**Sources of Funding**

The project was supported by Shanghai Municipal Key Clinical Specialty (grant number shslczdzk03202).

**Disclosures**

None

**Ethical approval**

Ethics committee of Zhongshan Hospital, Fudan University, Shanghai, China, gave ethical approval for this work.

**Table 1 Patient demographics. Total patient cohort (n =92)**

| Age  (years) | Mean ± SD | 61 ± 8 |
| --- | --- | --- |
|  | Median | 62 |
|  | Range | 35 – 75 |
| Weight (kg) | Mean ± SD | 68.3 ± 10.3 |
| Height (cm) | Mean ± SD | 166.7 ± 8.1 |
| Gender, n[%] | Male | 62 (67.4%) |
|  | Female | 30 (32.6%) |
| Lesion | LAD | 86 (61.0%) |
|  | LCX | 19 (13.5%) |
|  | RCA | 36 (25.5%) |
|  | Total | 141(100%) |
| No. of diseased vessels | 1 | 55 (59.1%) |
|  | 2 | 25 (26.9%) |
|  | 3 | 12 (12.9%) |
| Vessels with FFR | ≤0.8 | 38 (27.0%) |
|  | ＞0.8 | 103 (73.0%) |
|  | Total | 141(100%) |
| Cardiovascular risk factors, n (%) | | |
| hypertension | | 52 (56.5%) |
| hyperlipidemia | | 29 (31.5%) |
| diabetes | | 18 (19.6%) |
| Former or current smoker | | 25 (27.2%) |
| Family history of CAD | | 8 (8.7%) |
| Medication, n (%) | | |
| Platelet antagonist | | 17 (18.5%) |
| Lipid-lowering drugs | | 44 (47.8%) |
| Beta-blockers | | 19 (20.7%) |
| ARB/ACEI | | 28 (30.4%) |
| CCB | | 28 (30.4%) |
| Diuretics | | 6 (6.5%) |
| Insulin | | 4 (4.3%) |
| Orally antidiabetics | | 13 (14.1%) |
| Anticoagulant | | 35 (38.0%) |
| Anti-angina pectoris | | 11 (12.0%) |

LAD= left anterior descending; LCX = circumflex; RCA = right coronary artery.

**Table 2 Plaque characteristics and CMR data**

|  | | FFR Negative  （n = 103） | FFR Positive  （n = 38） | p-Value |
| --- | --- | --- | --- | --- |
| Lesion | LAD | 57 (66.3%) | 29 (33.7%) | 0.037 |
|  | LCx | 14 (73.7%) | 5 (26.3%) |  |
|  | RCA | 32 (88.9%) | 4 (11.1%) |  |
| Plaque Length  (mm) | | 27.2 ± 11.6 | 37.1 ± 15.3 | < 0.001 |
| MLA  (${mm}^{2}$) | | 3.4  (2.3 – 5.3) | 1.5  (0.9 – 2.1) | < 0.001 |
| Plaque Area  (${mm}^{2}$) | | 8.1 ± 3.3 | 10.1 ± 4.3 | 0.007 |
| PAS  (%) | | 53.4 ± 16.3 | 74.4 ± 15.3 | < 0.001 |
| TAV  (${mm}^{3}$) | | 171.6 ± 110.4 | 270.1 ± 164.2 | < 0.001 |
| Vessel Volume  (${mm}^{3}$) | | 360.5 ± 202.4 | 441.9 ± 235.7 | 0.045 |
| Lipid Rich Volume  (${mm}^{3}$) | | 10.3  (5.7 – 25.4) | 21.7  (9.0 – 51.6) | 0.010 |
| Lipid Plaque Ratio  (%) | | 11.6 ± 9.3 | 14.0 ± 11.5 | 0.204 |
| Spotty Calcium | | 23 (22.3%) | 21 (55.3%) | < 0.001 |
| Positive Remodeling | | 55 (53.4%) | 27 (71.1%) | 0.059 |
| Napkin-Ring Signs | | 13 (12.6%) | 15 (39.5%) | < 0.001 |
| Low attenuation Plaque | | 33 (32.0%) | 17 (44.7%) | 0.162 |
| Stress-MBF  (mL/(g·min)) | | 2.8 ± 0.7 | 2.4 ± 0.7 | 0.007 |
| MPR | | 2.6 ± 0.9 | 1.6 ± 0.4 | < 0.001 |

MLA = minimum luminal area; PAS = percent area stenosis; TAV = total atheroma volume; MBF = myocardial blood flow; MPR = myocardial perfusion ratio.

**Table 3 Univariate and multivariate logistic regression analysis of plaque characteristics**

| Variables | Univariate Analysis |  | Multivariate Analysis |  |
| --- | --- | --- | --- | --- |
|  | Odds Ratio  （95% CI） | p-Value | Odds Ratio  （95% CI） | p-Value |
| Plaque Length | 1.059  (1.027 – 1.093) | < 0.001 | -- | -- |
| MLA | 0.305  (0.188 – 0.496) | < 0.001 | 0.284  (0.166 – 0.485) | < 0.001 |
| Plaque Area | 1.142  (1.033 – 1.262) | 0.009 | -- | -- |
| PAS | 1.095  (1.058 – 1.134) | < 0.001 | -- | -- |
| TAV | 1.006  (1.002 – 1.009) | < 0.001 | 1.006  (1.002 – 1.010) | 0.003 |
| Vessel Volume | 1.002  (1.000-1.003) | 0.049 | -- | -- |
| Lipid Rich Volume | 1.016  (1.005 – 1.027) | 0.005 | -- | -- |
| Spotty Calcium | 0.233  (0.106 – 0.513) | < 0.001 | -- | -- |
| Napkin-Ring Signs | 0.221  (0.093 – 0.530) | 0.001 | -- | -- |

MLA= minimum luminal area; PAS= percent area stenosis; TAV= total atheroma volume.

**Table 4 Diagnostic performance of stress MBF, MPR, and Plaque Characteristic Index for detecting lesion-specific ischemia**

|  | AUC  (95% CI) | Best Cutoffs | Sensitivity  (95% CI)  (%) | Specificity  (95% CI)  (%) | PPV (%) | NPV (%) | Accuracy (%) |
| --- | --- | --- | --- | --- | --- | --- | --- |
| Stress MBF  (mL/(g·min)) | 0.638  (0.553- 0.718) | ≤ 2.61 | 55.3 (38.3- 71.4) | 63.1 (53.0- 72.4) | 35.6 (27.4- 44.7) | 79.3 (72.3- 84.9) | 61.0 (52.8- 69.1) |
| MPR | 0.921  (0.863- 0.959) | ≤ 1.82 | 89.5 (75.2- 97.1) | 89.3 (81.7- 94.5) | 75.6  (63.6- 84.5) | 95.8  (90.1- 98.3) | 89.4 (84.2- 94.5) |
| Plaque Characteristic Index | 0.885  (0.821- 0.933) | > 0.368 | 79.0 (62.7- 90.4) | 84.5 (76.0- 90.9) | 65.2  (53.7- 75.2) | 91.6  (85.4- 95.3) | 82.3 (75.9- 88.7) |

ROC = receiver operating characteristic; AUC = the area under the ROC curve; PPV = positive predictive value; NPV = negative predictive value; CI = confidence interval.

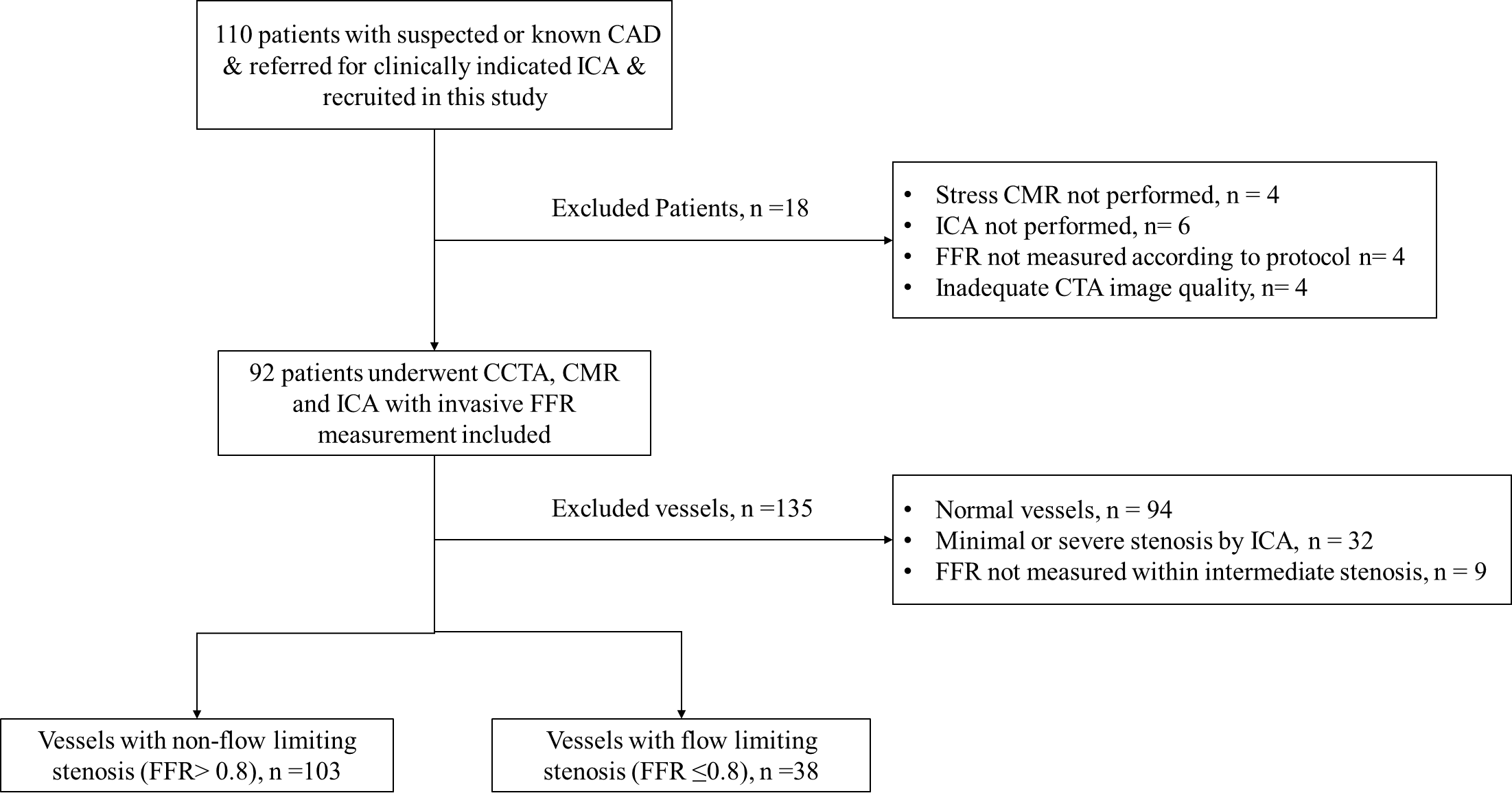

**Figure 1. Study flow chart**

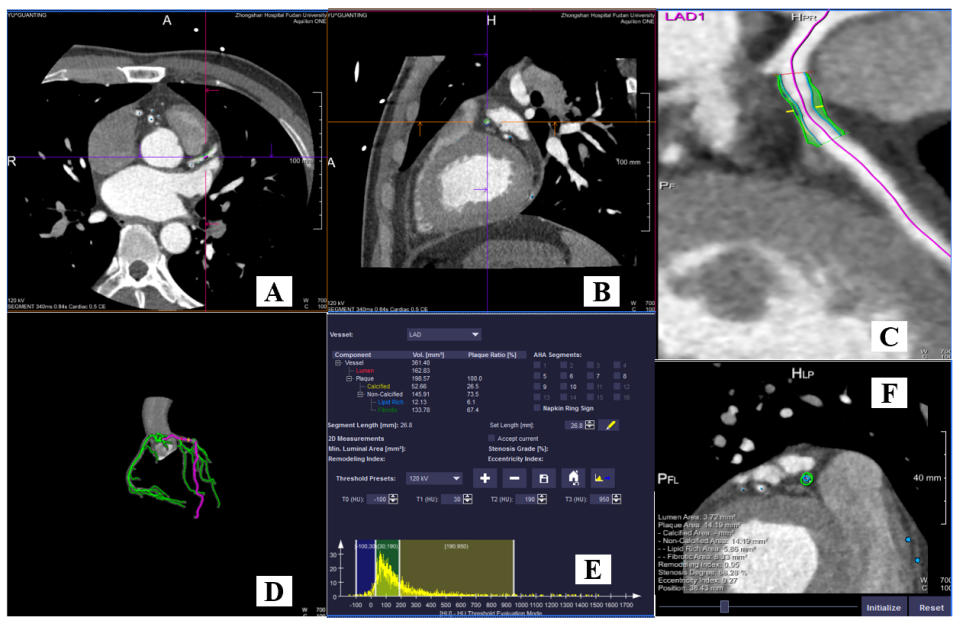

**Figure 2. Case example of CCTA-derived plaque characteristic index** **analysis.**

Representative patient who had underwent CCTA for suspected CAD. (A, B) The original image data of the patient. (C, D) Reconstruction image and model. (E) Color-coded semiautomatic plaque quantification of the lesion shows plaque length (18.9mm), plaque area (10.47${mm}^{2}$), MLA (2.92 ${mm}^{2}$) and etc. (F) 2D plaque characteristics analysis shows the lesion without any spotty calcium, positive remodeling, napkin-ring signs and low attenuation plaque.

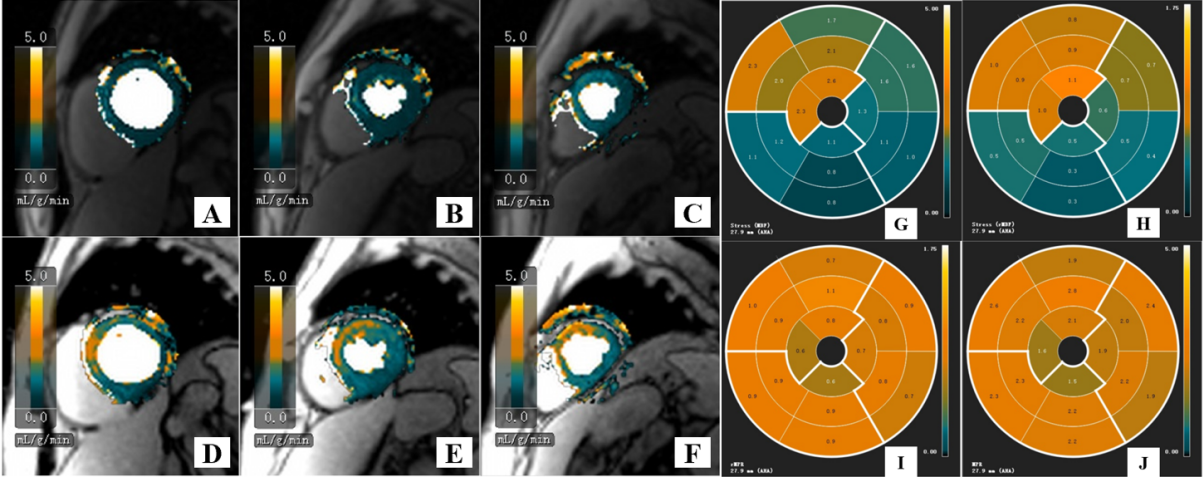

**Figure 3. Case example of quantitative CMR perfusion analysis.**

Representative patients with chest pain suspected of CAD. CCTA showed moderate stenosis of LAD. (A-E) Cardiac magnetic resonance (CMR) imaging of the basal left ventricular (A, D), midventricular (B, E), and apical (C, F) slices are shown during hyperemic (D-F) and resting (A-C) conditions. (G-J) The 17-segment Bulls-eye diagrams showing the distribution of CMR perfusion of stress MBF (G, H) and MPR (I, J) according to LAD.

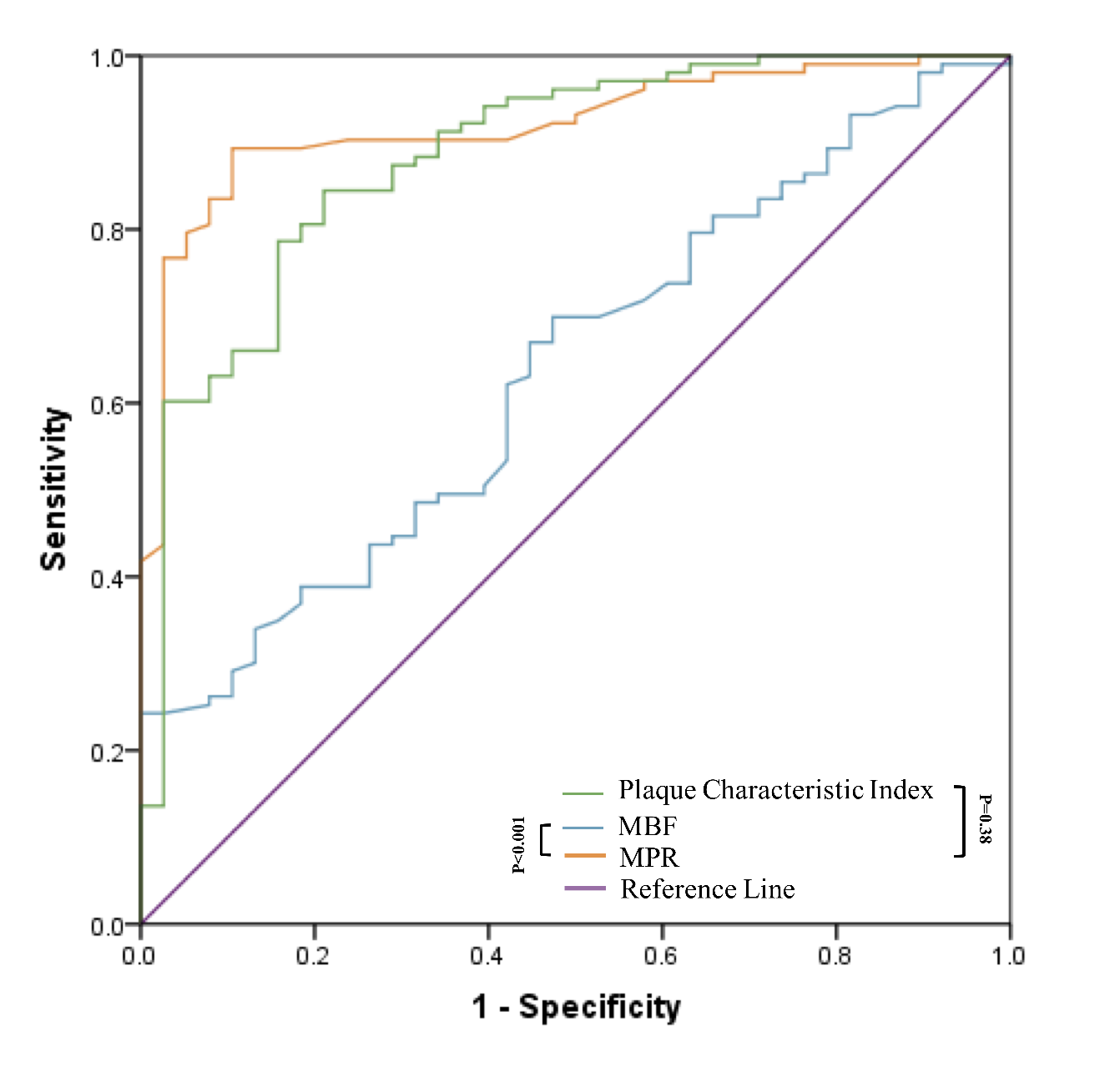

**Figure 4. Diagnostic performance of stress MBF, MPR, and Plaque Characteristic Index for the prediction of lesion-specific ischemia.**

The ROC analysis demonstrated that AUC of MPR (0.921, 95% CI: 0.863–0.959) and plaque characteristic index (0.885, 95% CI: 0.821–0.933, p=0.38) were higher than stress MBF (0.638, 95% CI: 0.553–0.718). MBF= myocardial blood flow; MPR= myocardial perfusion reserve.

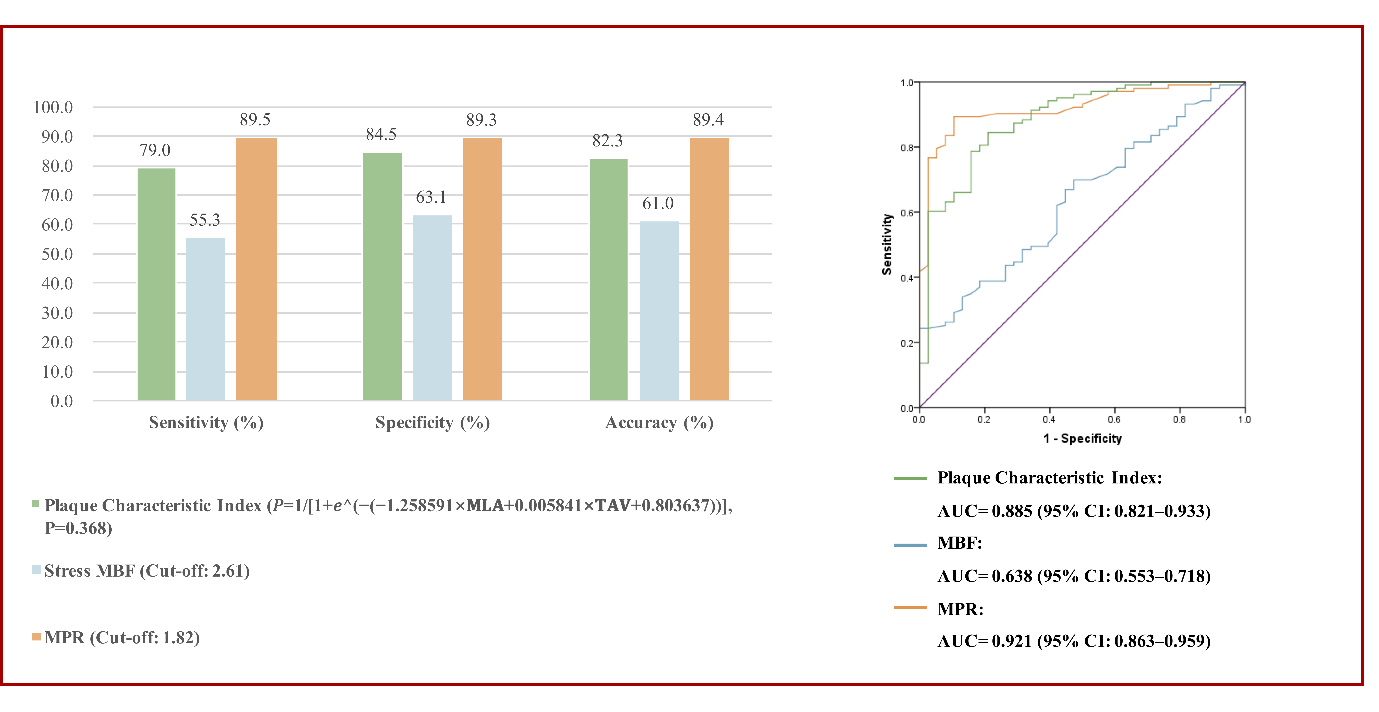

**Central illustration. Diagnostic Performance of Plaque Characteristic Index derived from Coronary Computed Tomography Angiography, Stress Myocardial Blood Flow and Myocardial Perfusion Reserve to Identify Hemodynamically Significant Stenosis in Patients with Stable Coronary Artery Disease.**

The receiver-operating characteristic curve and corresponding area under the curve describing the diagnostic performance of plaque characteristic index derived from Coronary Computed Tomography Angiography (CCTA), stress myocardial blood flow (MBF) and myocardial perfusion reserve (MPR) to identify hemodynamically significant stenosis as defined by invasive coronary angiography (ICA) with fractional flow reserve (FFR) at the vessel level.
