## SUPPLEMENTAL MATERIALS for "Comparison of CT-Derived Plaque Characteristic Index with CMR Perfusion for Ischemia Diagnosis in Stable CAD"

**Short title:** CCTA and MPR in myocardial ischemia

**Word count:** 4930

|  | AUC  (95% CI) | Optimal cut-off value | Sensitivity  (95% CI)  (%) | Specificity  (95% CI)  (%) | PPV (%) | NPV (%) | Accuracy (%) |
| --- | --- | --- | --- | --- | --- | --- | --- |
| MLA  (${mm}^{2}$) | 0.860 (0.791-0.913) | ≤ 2.25 | 84.2 (68.7- 94.0) | 75.7 (66.3- 83.6) | 56.1 (47.0- 64.9) | 92.9 (86.1- 96.5) | 78.0 |
| TAV  (${mm}^{3}$) | 0.707 (0.624- 0.780) | >161.72 | 81.6 (65.7- 92.3) | 55.3 (45.2- 65.1) | 40.3 (34.1- 46.7) | 89.1 (80.3- 94.2) | 62.4 |

**Table S1. Diagnostic performance of CCTA- derived minimum luminal area (MLA) and total atheroma volume (TAV) for lesion-specific ischemia assessment.**

ROC = receiver operating characteristic; AUC = the area under the ROC curve; PPV = positive predictive value; NPV = negative predictive value; CI= confidence interval
