## Supplementary material for "Comparison of CT-Derived Plaque Characteristic Index with CMR Perfusion for Ischemia Diagnosis in Stable CAD": Responses to Reviewer 2

**Responses to Reviewer #1**

Referee(s)' Comments to Author:

Reviewer #1

Comments to the Author

**General Comments**

The authors have to be commended on undertaking this interesting study.

**Major Comments:**

None.

**Specific Comments:**

**Introduction section**

**Question 1**: Paragraph 1, last sentence: please provide the latest AHA guidelines in place of reference 3 to support this sentence.
**Answer 1**: According to your suggestion, we added the latest AHA guidelines “Lawton JS, Tamis-Holland JE, Bangalore S, et al. 2021 acc/aha/scai guideline for coronary artery revascularization: a report of the american college of cardiology/american heart association joint committee on clinical practice guidelines. Circulation. 2022;145:e18-e114” in place of original reference 3.

**Question 2**: Paragraph 2, third sentence: CCTA-derived.....please provide reference to the CRESCENT Trial.

**Answer 2**: According to your suggestion, we added the “Lubbers M, Dedic A, Coenen A, et al. Calcium imaging and selective computed tomography angiography in comparison to functional testing for suspected coronary artery disease: the multicentre, randomized crescent trial. Eur Heart J. 2016;37:1232-43.” as the reference of “CCTA–derived quantitative plaque characteristic…” at paragraph 2.

**Question 3**: It is unclear if the authors wish to compare CCTA derived parameters with CMR perfusion parameter or these two with invasive FFR.

**Answer 3**: The aim of our study was to investigate the diagnostic performance of CCTA-derived plaque characteristics compared with myocardial blood flow (MBF) and myocardial perfusion reserve (MPR) derived from CMR perfusion using FFR as the reference, in the assessment of lesion-specific ischemia.

**Question 4**: Last paragraph, last sentence: what is the difference between qualitative and visual analysis?
**Answer 4**: It is a spelling mistake that we were intended to place the word “quantitative” at that place. Since we haven’t compared quantitative analysis of CMR perfusion images with visual analysis, we decided to delete this sentence (It needs to be emphasized that in our study, perfusion CMR is assessed quantitatively rather than visual analysis) from the paper. However, generally speaking, quantitative analysis of CMR perfusion images perform better than visual analysis, since it can give objective parameters and minimize interference from the analysts.(2)

**Methods section**: No comments

**Results**: What is the best cut-off value for minimal luminal area and TAV for ischemia that will be clinically relevant from the study?

**Answer:** The best cut-off value for MLA (minimal luminal area) and TAV (total atheroma volume) is 2.25 ${mm}^{2}$ and 161.72 ${mm}^{2}$,respectively. Detailed diagnostic accuracy statistics for MLA and TAV were shown at the following table.

|  | AUC  (95% CI) | Best Cutoffs | Sensitivity  (95% CI)  (%) | Specificity  (95% CI)  (%) | PPV (%) | NPV (%) |
| --- | --- | --- | --- | --- | --- | --- |
| MLA  (${mm}^{2}$) | 0.860 (0.791-0.913) | ≤ 2.25 | 84.2 (68.7- 94.0) | 75.7 (66.3-83.6) | 56.1 (47.0- 64.9) | 92.9 (86.1- 96.5) |
| TAV  (${mm}^{3}$) | 0.707 (0.624- 0.780) | ≤161.72 | 81.6 (65.7- 92.3) | 55.3 (45.2- 65.1) | 40.3 (34.1- 46.7) | 89.1 (80.3- 94.2) |

**Discussion**: No comments.
