## Supplementary material for "Comparison of CT-Derived Plaque Characteristic Index with CMR Perfusion for Ischemia Diagnosis in Stable CAD": Reply to the editor and reviewers

Dear Editor and Reviewers,

Thank you for your response letter. Our original manuscript titled “Comparison of CT-Derived Quantitative Plaque Characteristics with CMR Perfusion for Ischemia Diagnosis in Stable CAD” (CIRCCVIM/2023/015306) had been submitted to *Circulation: Cardiovascular Imaging.* Even though our work was not acceptable for publication in *Circulation: Cardiovascular Imaging* in its present form, we thank the Editor and reviewers for the time and effort that they have put into reviewing the previous version of the manuscript. Their suggestions have greatly inspired us and encouraged us to improve our work. And we tried our best to addresses the all of the comments of the referees, and decided to resubmit our manuscript.

Based on the instructions provided in your letter, we uploaded the file of the revised manuscript. Accordingly, we have uploaded a copy of the original manuscript with all the changes highlighted in MS Word.

Appended to this letter is our item-by-item response to the comments raised by the reviewers. The comments are reproduced and our responses are given directly afterward.

Thank you for allowing us to resubmit a revised copy of the manuscript. We hope that the revised manuscript can be accepted for publication in the *Circulation: Cardiovascular Imaging*.

Sincerely,

Mengsu Zeng, MD, PhD

Department of Radiology, Zhongshan Hospital,

Fudan University, Shanghai 200032, China
