## Supplementary material for "Comparison of CT-Derived Plaque Characteristic Index with CMR Perfusion for Ischemia Diagnosis in Stable CAD": Response to editor

Major comments

Question 1: On top of the concerns raised by the reviewers, the editorial team also requests a better definition of the included population, if possible with a clear flow chart. Were those patients initially candidates for invasive angiography, or were they selected only after invasive angiography? What was the criteria to enter the study and when was that defined (before ICA, after ICA, before or after CT / CMR). When was the CT and CMR performed? before or after invasive angiography? Please clarify the inclusion criteria and workflow for patient selection.

Answer 1: Thank you for your constructive question. We sorted out the whole study process based on collected patients’ information, clarified the included population and modified the study flow chart. A total of 110 patients with suspected or known CAD who were initially candidates for invasive coronary angiography were prospectively recruited. And the criteria to enter the study were defined before ICA, which means patients were selected before ICA. Additionally, CT and CMR were performed before ICA within 2 weeks. Detailed study design and study flow chart was shown in revised manuscript.
