## Supplementary material for "Comparison of CT-Derived Plaque Characteristic Index with CMR Perfusion for Ischemia Diagnosis in Stable CAD": Responses to Reviewer 1

Referee(s)' Comments to Author:

Reviewer #1

Comments to the Author

**General Comments**

Mengsu Zeng and coauthors from Zhongshan Hospital in Shanghai compare and analyze the associations and diagnostic performance of quantitative stress perfusion CMR myocardial blood flow (MBF) and perfusion reserve (MPR) as well as quantitative plaque characteristics derived from CCTA in a prospective, single center study that uses FFR as the reference standard. In brief, they find that CMR quantitative MBF did not perform as well as CMR MPR or a CCTA quantitative index combining minimum luminal area (MLA) and, as best I can tell, total atheroma volume (TAV). The diagnostic performance of CMR MPR and the CCTA quantitative index were both very good and not statistically different. Both were statistically superior to CMR MBF.

**Major Comments:**

**Question 1**: On major issue with this paper is the circular logic of developing a CCTA quantitative index and determining its diagnostic performance on the same patient dataset. The methods for describing exactly how the CCTA index was created need to be described in the methods. In the end, it appears that the index was derived from the multivariable regression results of the CCTA measurements of MLA and TAV. It also appears in the discussion that another paper has found those two parameters particularly good for diagnosing stable CAD.

**Answer 1:**

Based on our understanding of your question, we restated the logic of CCTA-derived plaque characteristics logistic regression model in the statistical analysis section. Parameters of CCTA-derived plaque characteristics that significantly differed between the flow-limiting group and non-flow-limiting group were then included into the univariate and subsequent forward stepwise multivariate logistic regression analysis. CCTA-derived plaque characteristics logistic regression model was created based on the independent predictors of lesion-specific ischemia.

**Question 2**: Since it is better to create and validate analysis models on different datasets, the authors need to add this as a limitation of the study. In light of the exploratory and developmental nature of the paper, the authors should add diagnostic accuracy statistics for CCTA MLA and TAV, the individual components from the CCTA that they found had statistically significant in the multivariable model. These could be based on the Youden index for MLA and TAV, a methodology at least similar to how they dealt with the CMR MBF and MPR. They should add the derived formula providing the relative weighting of MLA and TAV in their combined index. They could show the diagnostic accuracy statistics for the combined index (which are already in the paper).

**Answer 2:** Thanks for your suggestion, we add “Finally, the CCTA-Derived plaque characteristic index needs to validate on a new dataset” as our study limitation. Based on our understanding of your question, we supplemented a following table about the respective diagnostic accuracy statistics for MLA and TAV (derived from CCTA) and we put it into supplementary materials. The relative weighting of MLA and TAV in the Multivariate logistic regression of lesion-specific ischemia were -1.25891 and 0.005841, respectively (P<0.001, P=0.003, respectively), and the regression equation for predicting probability P was $P=1/[1+e^{-(-1.258591\times MLA+0.005841\times TAV+0.803637)}]$.

|  | AUC  (95% CI) | Best Cutoffs | Sensitivity  (95% CI)  (%) | Specificity  (95% CI)  (%) | PPV (%) | NPV (%) |
| --- | --- | --- | --- | --- | --- | --- |
| MLA  (${mm}^{2}$) | 0.860 (0.791-0.913) | ≤ 2.25 | 84.2 (68.7- 94.0) | 75.7 (66.3-83.6) | 56.1 (47.0- 64.9) | 92.9 (86.1- 96.5) |
| TAV  (${mm}^{3}$) | 0.707 (0.624- 0.780) | ≤161.72 | 81.6 (65.7- 92.3) | 55.3 (45.2- 65.1) | 40.3 (34.1- 46.7) | 89.1 (80.3- 94.2) |

**Question 3**: A second significant limitation is the description of the number of patients and number of vessels analyzed. Figure 3 indicates that out of 110 patients, 183 vessels were analyzed. One would have expected 330 (LAD, Cx, RCA) or 440 vessels (LM, LAD, Cx, RCA). Perhaps you only list vessels determined to be in the 30-90% stenosis range. Please clarify this in the methods and results sections. The results text should acknowledge that 41 vessels were excluded due to CCTA image quality. This should also be added to the limitations section.

**Answer 3**: It’s true that only patients with 30% to 90% stenosis of ≥1 major epicardial vessels determined by ICA were included in our study, and we have clarified it in the methods and results sections. Thanks to your comment, we found that there were some mistakes with the description of patients exclusion process, as the 41 vessels excluded were not just due to image quality. Of 110 patients enrolled, 106 patients underwent all noninvasive imaging within 2 weeks before ICA, and 14 patients were excluded due to not undergo CMR (n=4) or ICA (n=6) or invasive FFR (n=4). The other 4 patients were excluded for inadequate CCTA image quality. Therefore, 92 patients comprised the final study population, of whom 141 vessels with stenosis ranged between 30% and 90% underwent all tests. Moreover, we have added “A total of 18 patients were excluded, which may incur a selection bias” as one of our limitations.

**Specific Comments:**

**Question 1**: Diagnostic accuracy statistics should be reported to on significant digit in the abstract, paper, tables, and figures.

**Answer 1**: According to your advice, diagnostic accuracy statistics in our manuscript were reported to significant digit at three decimal places in the abstract, paper, tables, and figure.

**Question 2**: Page 9, Reference 21 is a more general reference to Fermi function deconvolution. The authors used a product from Circle CVi42 that was based on the methods of Hsu et al, JACC Cardiovasc Imaging. 2018 May;11(5):697-707. This method uses a modified Fermi function deconvolution.

**Answer 2**: We have added “Hsu LY, Jacobs M, Benovoy M, et al. Diagnostic performance of fully automated pixel-wise quantitative myocardial perfusion imaging by cardiovascular magnetic resonance. Jacc Cardiovasc Imaging. 2018;11:697-707.” as the reference of the Fermi-constrained deconvolution used in our study.

**Question 3**: Page 11. What was the reason one vessel was not eligible for FFR? How did the authors deal with vessels >90% stenosis?

**Answer 3**: The four patients were not eligible for FFR measurement and excluded due to extremely tortuous, calcified coronary vessels or chronic total occlusion. And for the patients with vessels with >90% stenosis, direct revascularization was performed immediately for them at the interventionalists discretion.

**Question 4**: Page 12. Add to the methods how you defined angina pectoris type I and type II.

**Answer 4**: Angina pectoris is one of the clinical symptoms, used for screening patients, and we decided not to distinguish different type of angina pectoris in the manuscript. As for the definition, angina pectoris type I is referred to typical angina, describing retrosternal chest discomfort, which usually happens during exertion and can be relieved by rest or nitroglycerin. Angina pectoris type II is referred to atypical angina, meeting two out of three criteria for typical angina pectoris, according to the reference below(1).

**Question 5**: Results - try to minimize repeating many numerical results that are also in the tables or figures. For example, putting confidence intervals in the results text is more difficult to read than in the tables or figures. Make sure pointers to the correct tables and figures accompany each result or result paragraph.

**Answer** **5**: According to your suggestion, we removed the redundant confidence intervals from the result text, only retaining them in the tables and figures. We modified our tables and figures in order to ensure each result or result paragraph accompanied by the correct tables and figures.

**Question** **6**: Results, pg 13, end of first paragraph. Add the word "independent" between "significant" and "predictors".

**Answer 6**: We have added the word "independent" and marked it in yellow at the place your mentioned.

**Question 7**: Page 14. Avoid changing the terminology of your CCTA model that combines MLA and TAV. Pick one name and use that throughout the paper to minimize reader confusion.

**Answer 7**: Thanks for your suggestion. To minimize reader confusion, we chose “CCTA-derived plaque characteristic Index” as the terminology of our multivariate logistic regression model in the whole paper.

**Question 8**: Discussion, page 15, first full paragraph. The abbreviation "PAV" appears twice. Did you mean "TAV"?

**Answer 8**: “PAV” isn’t a spelling mistake and is referred to “percent atheroma volume”. In our first version of manuscript, we used its abbreviation “PAV” when we first mentioned it in the paper, leading to a misunderstanding. We have changed the expression and marked it in yellow in our new version of manuscript.

**Question 9**: Central Figure. Add some of the diagnostic accuracy statistics.

**Answer 9**: According to your suggestion, we added the diagnostic accuracy statistics and remade our central figure.

**Grammatical suggestions:**

Pg 5, Introduction, first sentence. "Coronary artery disease (CAD) is one of the major cardiovascular diseases affecting the global human population and has been proven to be the major cause of death(1)." Consider rewriting this sentence as follows: "Coronary artery disease (CAD) is a global problem and a leading cause of death(1)."

Pg 5, Introduction, second sentence. Rewrite the sentence which currently describes ischemic wall motion abnormalities as "irreversible". While myocardial infarction can cause permanent regional wall motion abnormalities, ischemia can occur in the absence of infarction and a significant subset of non-infarcted regional wall motion abnormalities show recovery of function after successful revascularization.

Pg 5, Introduction, third sentence. Consider changing "instructing" to "guiding"

Pg 5, Introduction, 2nd paragraph, 1st sentence. Consider changing "due to" to "since it is".

Pg 5, Introduction, 2nd paragraph. Consider revising "high specificity and negative predictive value for diagnosing obstructive CAD" to "high sensitivity and negative predictive value for diagnosing obstructive CAD".

Pg 5, Introduction, 2nd paragraph. Consider changing "perfoming" to "and performed".

Pg 5, Introduction, 2nd paragraph. Consider changing "was proven" to "has"

Pg 7, methods, 2nd paragraph. Consider changing "As for diffused lesions..." to "As for diffuse lesions..."

Pg 7, methods, 2nd paragraph. Consider deleting the word "immediately" from "...were measured immediately as a reference..."

Pg 8, methods, first sentence. Consider changing "length/area" to "length or area". Also consider deleting "... that plaque located."

Pg 11, Results, 2nd sentence. Did the exclusion due to not being eligible for FFR include stenoses >90%? Also, breakdown FFR exclusions (1) and CCTA exclusions (41) in the results text. This is an important result.

Pg 13, Results, last paragraph. Consider changing "...while both of them were significantly higher..." to "...while both were significantly higher..."

Pg 16. Discussion, 2nd sentence, starting with "Additionally, We also found..." Please divide this sentence into 2 or 3 shorter sentences.

**Answer to grammatical suggestions:** Thanks for your advice on our grammar improvement. We have adjusted the corresponding word or sentence and marked them in yellow in the paper.
